# Non-invasive Detection of Fasciculation Using Surface EMG with a Wavelet-Based Analytical Method (DEWCS)

**DOI:** 10.64898/2026.06.15.26355644

**Authors:** Takahiko Mukaino, Hidetoshi Nagai, Yuko Kobayakawa, Senri Ko, Kazunori Iwao, Kotaro Iida, Takashi Irie, Saeko Inamizu, Satoshi Nagata, Eizo Tanaka, Ryo Kurasawa, Hajime Takeuchi, Eri Miyazaki, Noriko Isobe, Hiroshi Shigeto

## Abstract

**Objective:** Needle electromyography (nEMG) is essential for diagnosing neuromuscular disorders but is invasive and often painful. We employed single-channel bipolar surface EMG (sEMG) analyzed with a novel wavelet-based analytical approach, “Detecting and Extracting Elemental Wave Components based on a Wavelet Coefficient Set (DEWCS)” and investigated whether fasciculation-related activity could be identified.

**Methods:** In this prospective study, 28 patients undergoing nEMG for suspected neuromuscular disorders and 13 healthy controls were included. Resting-state sEMG was recorded from selected muscles using single-channel bipolar active electrodes at a high sampling rate. DEWCS was used to extract indices reflecting fast- and slow-type motor unit (MU)-related activity. These standardized indices were evaluated against nEMG-detected fasciculation potentials using generalized estimating equation logistic regression to account for within-subject clustering. Diagnostic performance was assessed by receiver operating characteristic analysis.

**Results:** A total of 67 muscles from 38 participants were analyzed. Indices of fast- and slow-type MU-related activity were significantly associated with fasciculation potentials (slow: OR 5.10, p = 0.0041; fast: OR 2.38, p = 0.0162). The combined model showed excellent discrimination (area under the curve = 0.97), outperforming either index alone. Muscle region had no significant effect.

**Conclusions:** A single-channel bipolar sEMG setup combined with DEWCS detected fasciculation-related activity with promising accuracy. This method may serve as a non-invasive surrogate marker of lower motor neuron involvement. Further validation in larger cohorts is warranted.

**Significance:** This non-invasive sEMG approach may help detect fasciculation-related activity and complement nEMG in neuromuscular diagnostics.

## 1. Introduction

Needle electromyography (nEMG) is an essential examination for the diagnosis of neuromuscular disorders. In addition to distinguishing neurogenic from myogenic disorders, it plays a crucial role in assessing ongoing denervation and abnormal spontaneous activity. However, nEMG is painful and invasive, and the procedure is burdensome not only for patients but also for the physicians who perform it (London, 2017).

Surface electromyography (sEMG) is a non-invasive technique for recording muscle electrical activity, and several studies have explored its use in the diagnosis of neuromuscular disorders. For example, recordings from the tibialis anterior (TA) have been reported to differentiate neurogenic from myogenic processes (Uesugi et al., 2011), and high-density sEMG has been used to detect or quantify fasciculation potentials, thereby potentially aiding in the diagnosis of amyotrophic lateral sclerosis (Bashford et al., 2020). Nevertheless, to date, despite promising results in research settings, its clinical adoption remains limited, likely due to practical challenges in electrode setup, signal processing, and the lack of standardized workflows.

Recently, Nagai developed an analytical method to estimate motor unit–related activity from sEMG signals using redundant wavelet coefficient analysis (Nagai, 2023b). This method enables extraction of motor unit action potential-like components from sEMG recordings obtained at 10–20 kHz. We conducted a prospective study to determine whether this technique could identify fasciculation-related activity at rest, using fasciculation potentials on nEMG as the reference standard. In contrast to high-density approaches, the present method uses a simple single-channel recording, which may facilitate its application in routine clinical settings.

## 2. Methods

This prospective study was conducted in the electrophysiology laboratory at Kyushu University Hospital between April 2025 and March 2026. The institutional review board of Kyushu University Hospital approved this study (approval number: 24003), and the study was registered in the Japan Registry of Clinical Trials (jRCT1072240102). Written informed consent was obtained from all participants. No treatment allocation or modification of clinical management was performed.

### 2.1 Participants

We enrolled patients aged ≥18 years who were suspected of having neuromuscular disorders and underwent nEMG, as well as healthy volunteers with no history of neuromuscular disease. Individuals in whom nEMG was anticipated to be difficult—such as those with marked sensitivity to pain or inability to remain still, those with extensive skin lesions at the recording site, and those otherwise deemed unsuitable—were excluded. Based on the clinical diagnosis at the time of analysis, participants were categorized into five groups: Myogenic, Neurogenic-LMN (neurogenic disorders with lower motor neuron involvement), Neurogenic-UMN (neurogenic disorders with upper motor neuron involvement without lower motor neuron involvement), Normal (no neurogenic or myogenic disorder, including healthy controls), and Undetermined.

### 2.2 EMG recording protocol

For each patient, routine nEMG was first performed for diagnostic purposes by the examiners (S.K., K.Iw., K.Ii., T.I., S.I., S.N., E.T., R.K., H.T., E.M.). The presence or absence of abnormal spontaneous activity and fasciculation potentials was independently evaluated by the examiners as part of the clinical assessment, and the final interpretation was used for analysis. After completing the nEMG examination, sEMG recordings were obtained from muscles selected based on the nEMG findings, prioritizing those with the most prominent abnormalities, or at least one muscle when no definite abnormality was identified.

The primary target muscles included the biceps brachii (BB), first dorsal interosseous (FDI), and tibialis anterior (TA), which were selected due to their clinical relevance and accessibility for surface recordings. Other muscles were also examined as appropriate, including the triceps brachii, vastus medialis, vastus lateralis, and rectus femoris.

### 2.3 sEMG acquisition

sEMG was recorded using custom-made active Ag/AgCl electrodes with a built-in amplification factor of 1,000 and a band-pass filter of 40–10,000 Hz. Signals were acquired at a high sampling rate (20,000 Hz; typical clinical sEMG: 1,000–2,000 Hz), after additional amplification (×1–14) by a post-amplifier.

For each target muscle, recordings during slight and forceful contractions and a 60-second resting-state recording were obtained; only the resting-state data were used for the present analyses. All recordings were semi-automated and guided by computer-generated audio instructions, and the examiners monitored electrode placement quality and recording stability throughout the procedure. sEMG examinations in both patients and healthy volunteers were performed by T.M. and Y.K. After visual inspection of the raw recordings, datasets with excessive noise or poor electrode contact (e.g., insufficient adhesion or detachment during recording) were excluded.

### 2.4 Data processing with the DEWCS method

sEMG signals were processed based on a previously described method by Nagai (Nagai, 2023a, 2023b), which enables detection of motor unit (MU)–related activity using redundant wavelet coefficient analysis applied to high-sampling-rate data. We refer to this framework as “Detecting and Extracting Elemental Wave Components based on a Wavelet Coefficient Set (DEWCS)”.

The DEWCS procedure is summarized as follows.

i. Raw sEMG signals were analyzed using a redundant discrete wavelet transform, in which wavelet coefficients were computed at every sampling time without downsampling, enabling a shift-invariant time–frequency representation of the signal.
ii. For each sampling time, a wavelet coefficient set (WCS) was constructed by collecting coefficients across multiple frequency bands according to predefined relative temporal offsets corresponding to the expected structure of motor unit action potentials (MUAPs). The WCS characterizes the waveform structure of MU-related activity (Fig. 1a, b).
iii. To evaluate whether a candidate WCS corresponded to a physiologically plausible MUAP, a matching ratio (MR) was calculated based on similarity to predefined reference vectors representing typical MUAP waveform characteristics. This evaluation allows for variations in waveform shape arising from differences in MU type or physiological conditions, such that waveforms falling within an acceptable range of variation are assigned similarly high matching scores (Fig. 1c).
iv. Each WCS was further allocated to slow- and fast-type MU components using a time–frequency allocation procedure. In this procedure, the MR-based contribution of each MU component was modulated by component-specific frequency weighting, so that lower-frequency coefficients were preferentially assigned to the slow-type MU component and higher-frequency coefficients to across *L* frequency levels, *W =*(w_-1_, ,w_-*L*_), the wavelet set strength (WSS) the fast-type MU component. For a WCS consisting of wavelet coefficients was calculated as *L* times the root-mean-square (RMS) of the coefficient values:

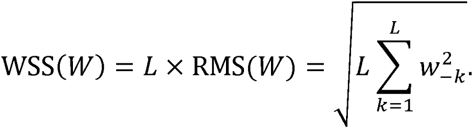 Thus, WSS represents a scaled RMS-based measure of WCS strength across frequency levels, emphasizing large coefficient values rather than simply summing the coefficients. WSS was calculated separately for the slow- and fast-type MU components after time–frequency allocation (Fig. 1d; Supplementary Fig. 1).
v. In the original DEWCS framework, MU-related activity was detected by identifying local-maxima in the product of MR and WSS (MR × WSS), which incorporates both waveform similarity and WCS strength. In the present study, the ranked distribution of MR × WSS values were used to derive the resting-state motor unit activity index (RS-MUAI), as described below.

**Figure 1.**
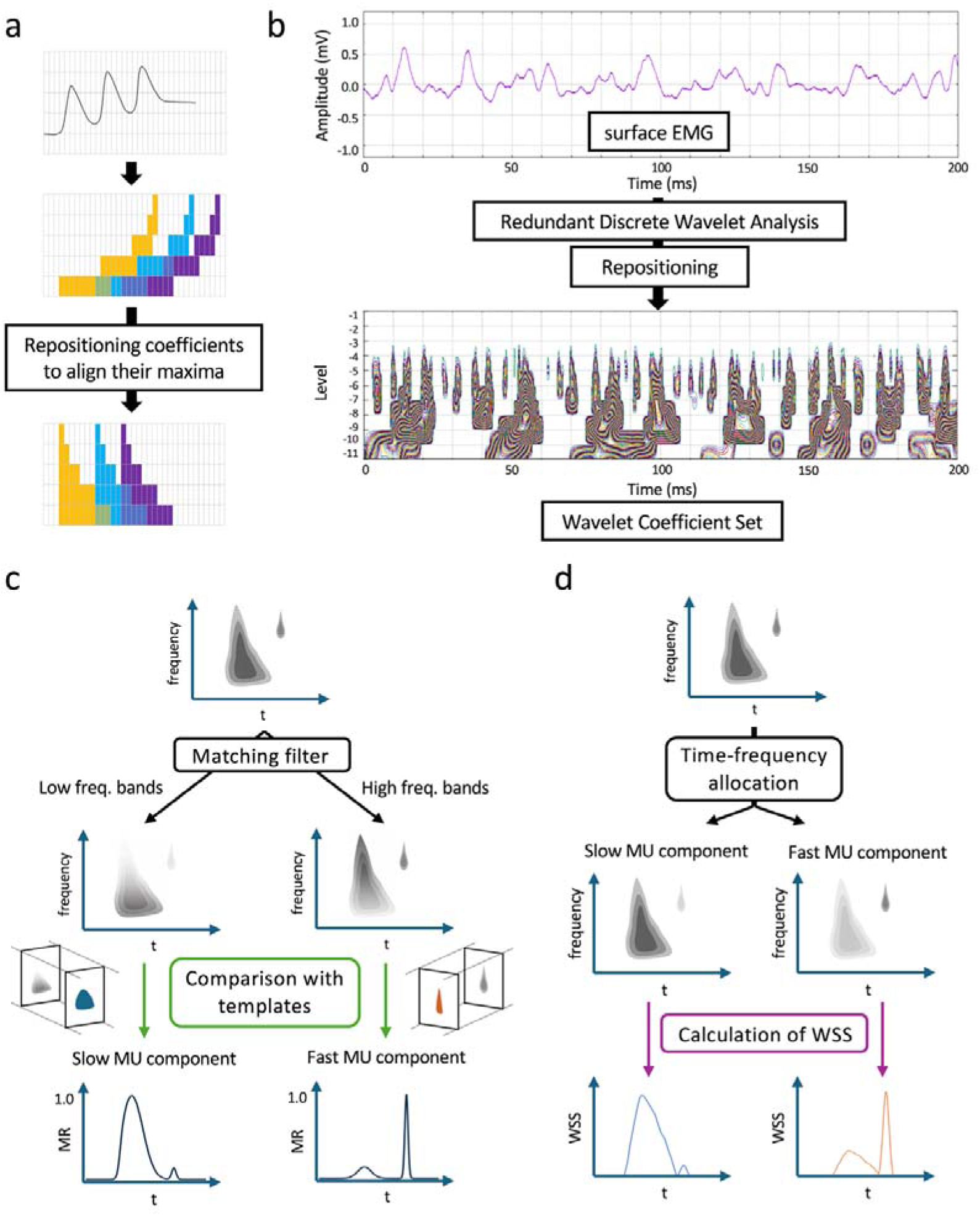
Schematic illustration of the DEWCS-based extraction of slow- and fast-type motor unit activity indices from surface electromyography. (a) A motor unit action potential (MUAP), an elemental waveform constituting the surface electromyography signal, is transient and non-periodic. Redundant discrete wavelet analysis was used to calculate wavelet coefficients at every sample time point. Because the coefficient maxima associated with a single MUAP appear at different relative times across frequency bands, the coefficients characterizing the waveform were repositioned so that their maxima were aligned along a line perpendicular to the time axis, thereby forming a wavelet coefficient set (WCS). (b) WCS candidates were extracted from the surface electromyography signal using redundant discrete wavelet analysis. Localized droplet-like coefficient structures represented candidate MUAPs. (c) Each WCS was processed using a matching filter that emphasized frequency-band patterns relevant to slow- and fast-type motor unit (MU) components. The filtered WCS was then compared with predefined templates for each MU component, and the degree of MUAP-likeness was quantified as the matching ratio (MR). (d) Each WCS was further allocated to slow- and fast-type MU components by time–frequency allocation, and the wavelet set strength (WSS) was then calculated separately for the slow- and fast-type MU components.

### 2.5 Application of the DEWCS method to resting-state sEMG

The DEWCS method was originally developed for analyzing sEMG during muscle contraction in healthy subjects, particularly to evaluate fast- and slow-type MU activity. For application to patients with neuromuscular disorders, initial parameter adjustment was performed by H.N. using a small subset of recordings from patients with clear neurogenic changes on nEMG and from healthy controls. During this parameter-setting phase, nEMG findings were available to the analyst. After the analytic parameters had been fixed, the same pipeline was applied to the full dataset without further modification.

Details of the application of the DEWCS method to resting-state sEMG have been described elsewhere (Nagai, 2026). In the present study, a modified approach based on this framework was used to quantify spontaneous MU-related activity at rest.

All candidate WCS events were ranked according to their MR × WSS values, and the distribution of these ranks was transformed using a logarithmic scale (Fig. 2a, b). This transformation compresses the long-tailed distribution corresponding to low-intensity and low-matching noise-like signals, while relatively expanding the distribution of high-intensity, high-matching candidates. The rank axis was normalized such that the median rank corresponded to 1.0, enabling comparison of distribution shapes across recordings.

**Figure 2.**
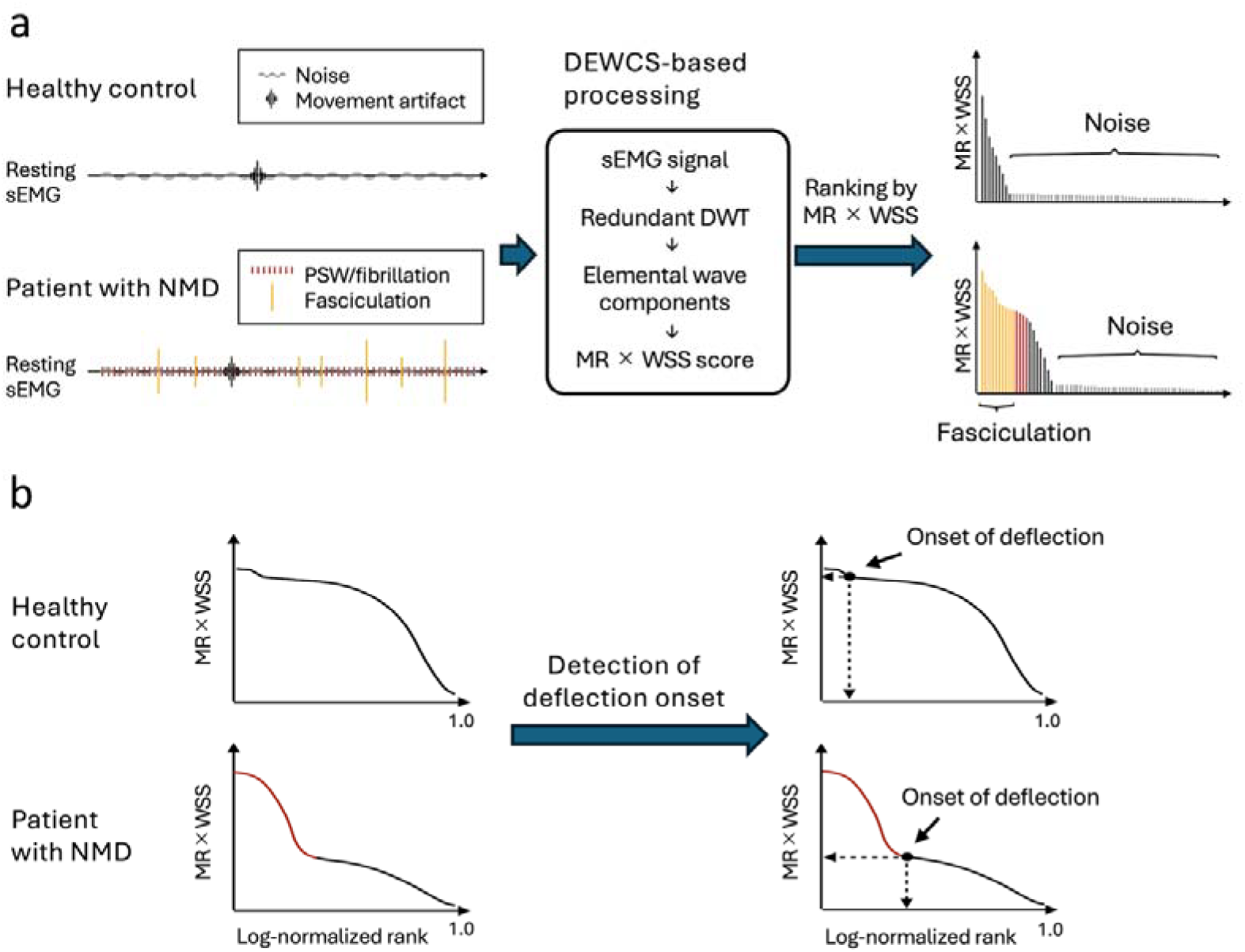
Schematic illustration of DEWCS-based processing of resting-state sEMG and derivation of RS-MUAI. (a) Representative resting-state surface electromyography (sEMG) signals are shown for a healthy control and a patient with a neuromuscular disorder (NMD). In healthy controls, the resting signal primarily reflects background noise, with occasional movement artifacts. In patients with NMD, abnormal spontaneous activity, particularly fasciculation-related motor unit activity, is superimposed on the resting signal, together with other components potentially related to positive sharp waves (PSWs) or fibrillation potentials. The sEMG signal is processed using the DEWCS-based algorithm, which applies redundant discrete wavelet transform, extracts elemental wave components, and calculates the MR × WSS score. The extracted components are then ranked in descending order according to MR × WSS. In healthy controls, the ranked profile is dominated by noise-like components, whereas in patients with NMD, high-ranking components corresponding to abnormal spontaneous motor unit activity, including fasciculation-related activity, appear before the noise-dominant portion. (b) MR × WSS values were arranged in descending order and plotted against the logarithmically normalized rank. In healthy controls, the profile gradually declines and is dominated by noise-like components. In patients with neuromuscular disorders (NMD), high-MR × WSS components related to abnormal spontaneous motor unit activity extend over a broader high-ranking range, resulting in a rightward shift of the deflection onset. The onset of deflection was identified, and its rank position was used to derive the RS-MUAI. The corresponding MR × WSS level can be interpreted conceptually as a threshold separating abnormal motor unit activity from noise-like components. Higher RS-MUAI values indicate a greater degree of abnormal spontaneous motor unit activity in resting-state sEMG.

In this log-rank representation, recordings unlikely to contain MU-related activity typically exhibit a smoothly varying curve across ranks, reflecting a homogeneous population of noise-like signals. In contrast, recordings containing spontaneous MU-related activity show a deviation from this smooth curve at higher ranks. The smoothly varying portion was interpreted as a distribution of noise-like components, whereas the deviating portion was interpreted as a subset of candidate events with distinct characteristics, presumed to correspond to spontaneous MU-related activity. The onset of this deviation was identified from the ranked MR × WSS distribution. In the original framework, the corresponding MR × WSS level can be interpreted as a threshold for classifying candidate events. In the present analysis, however, this amplitude threshold was not used for event classification; instead, the rank position at which the deviation began was used to derive the RS-MUAI. RS-MUAI increases when a larger subset of high-ranking events deviates from the noise-like distribution, thereby reflecting the relative prominence of spontaneous MU-related discharges at rest.

### 2.6 Statistical analysis

Using each muscle as a unit of analysis and nEMG findings as the reference standard, we examined whether RS-MUAIs reflecting fast- and slow-type MU-related activity (RS-MUAI (fast) and RS-MUAI (slow)) derived from resting-state sEMG could distinguish muscles with fasciculation potentials on nEMG from those without. Muscles were categorized into two groups based on nEMG findings: Fas-positive (presence of fasciculation potentials) and Fas-negative (absence of fasciculation potentials). The wavelet-based indices were calculated for each 60-second resting sEMG recording.

Because multiple muscles were assessed in some participants, observations at the muscle level were not independent. To account for within-subject correlation, generalized estimating equation (GEE) logistic regression models were used as the primary analytical approach, with subject ID specified as the clustering variable and an exchangeable working correlation structure.

In the primary model, the presence or absence of fasciculation potentials was treated as the dependent variable, and standardized resting-state motor unit activity indices (RS-MUAI (fast) and RS-MUAI (slow)) were included as independent variables. These indices were standardized using z-score transformation prior to analysis. In a secondary model, muscle region (upper vs. lower limb) was additionally included to assess its potential confounding effect.

Diagnostic performance was evaluated using receiver operating characteristic (ROC) analysis at the muscle level. Confidence intervals for the areas under the ROC curves (AUCs) were calculated using DeLong’s method. The optimal cut-off value was determined using the Youden index. Continuous variables are presented as median [interquartile range, IQR] or mean ± standard deviation, as appropriate. Group differences were evaluated using the Mann–Whitney U test or the Kruskal–Wallis test, as appropriate. A p-value < 0.05 was considered statistically significant. All statistical analyses were performed using R (version 4.5.2).

## 3. Results

The study included 28 patients with suspected neuromuscular disorders and 13 healthy volunteers. sEMG signals were initially obtained from 41 muscles in patients and 36 muscles in controls. Ten muscle recordings from three patients were excluded due to poor electrode attachment or excessive noise. Thus, 67 muscles from 38 participants, including 25 patients and 13 healthy volunteers, were included in the final analysis. In the patient group, nEMG identified fasciculation potentials in 11 muscles from 8 patients. Demographic and clinical characteristics of the participants are summarized in Table 1. There were significant differences among groups in sex distribution and age (p = 0.0442 and p < 0.0001, respectively), whereas height and weight did not differ significantly among groups. Detailed needle EMG findings and corresponding RS-MUAI values for the fasciculation-positive muscles are shown in Table 2.

**Table 1.**
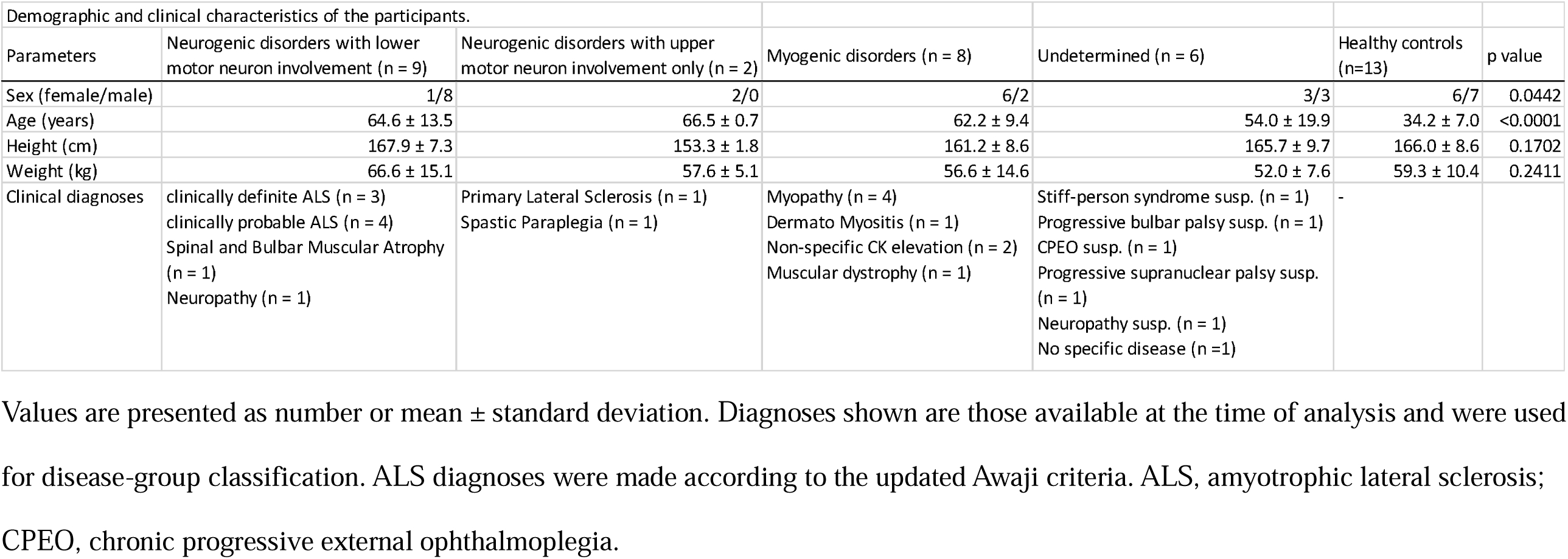
Demographic and clinical characteristics of the participants.

| Demographic and clinical characteristics of the participants. |  |  |  |  |  |  |
| --- | --- | --- | --- | --- | --- | --- |
| Parameters | Neurogenic disorders with lower motor neuron involvement (n = 9) | Neurogenic disorders with upper motor neuron involvement only (n = 2) | Myogenic disorders (n = 8) | Undetermined (n = 6) | Healthy controls (n=13) | p value |
| Sex (female/male) | 1/8 | 2/0 | 6/2 | 3/3 | 6/7 | 0.0442 |
| Age (years) | 64.6 ± 13.5 | 66.5 ± 0.7 | 62.2 ± 9.4 | 54.0 ± 19.9 | 34.2 ± 7.0 | <0.0001 |
| Height (cm) | 167.9 ± 7.3 | 153.3 ± 1.8 | 161.2 ± 8.6 | 165.7 ± 9.7 | 166.0 ± 8.6 | 0.1702 |
| Weight (kg) | 66.6 ± 15.1 | 57.6 ± 5.1 | 56.6 ± 14.6 | 52.0 ± 7.6 | 59.3 ± 10.4 | 0.2411 |
| Clinical diagnoses | clinically definite ALS (n = 3)<br>clinically probable ALS (n = 4)<br>Spinal and Bulbar Muscular Atrophy (n = 1)<br>Neuropathy (n = 1) | Primary Lateral Sclerosis (n = 1)<br>Spastic Paraplegia (n = 1) | Myopathy (n = 4)<br>Dermato Myositis (n = 1)<br>Non-specific CK elevation (n = 2)<br>Muscular dystrophy (n = 1) | Stiff-person syndrome susp. (n = 1)<br>Progressive bulbar palsy susp. (n = 1)<br>CPEO susp. (n = 1)<br>Progressive supranuclear palsy susp. (n = 1)<br>Neuropathy susp. (n = 1)<br>No specific disease (n =1) | - |  |

Values are presented as number or mean ± standard deviation. Diagnoses shown are those available at the time of analysis and were used for disease-group classification. ALS diagnoses were made according to the updated Awaji criteria. ALS, amyotrophic lateral sclerosis; CPEO, chronic progressive external ophthalmoplegia.

**Table 2.**
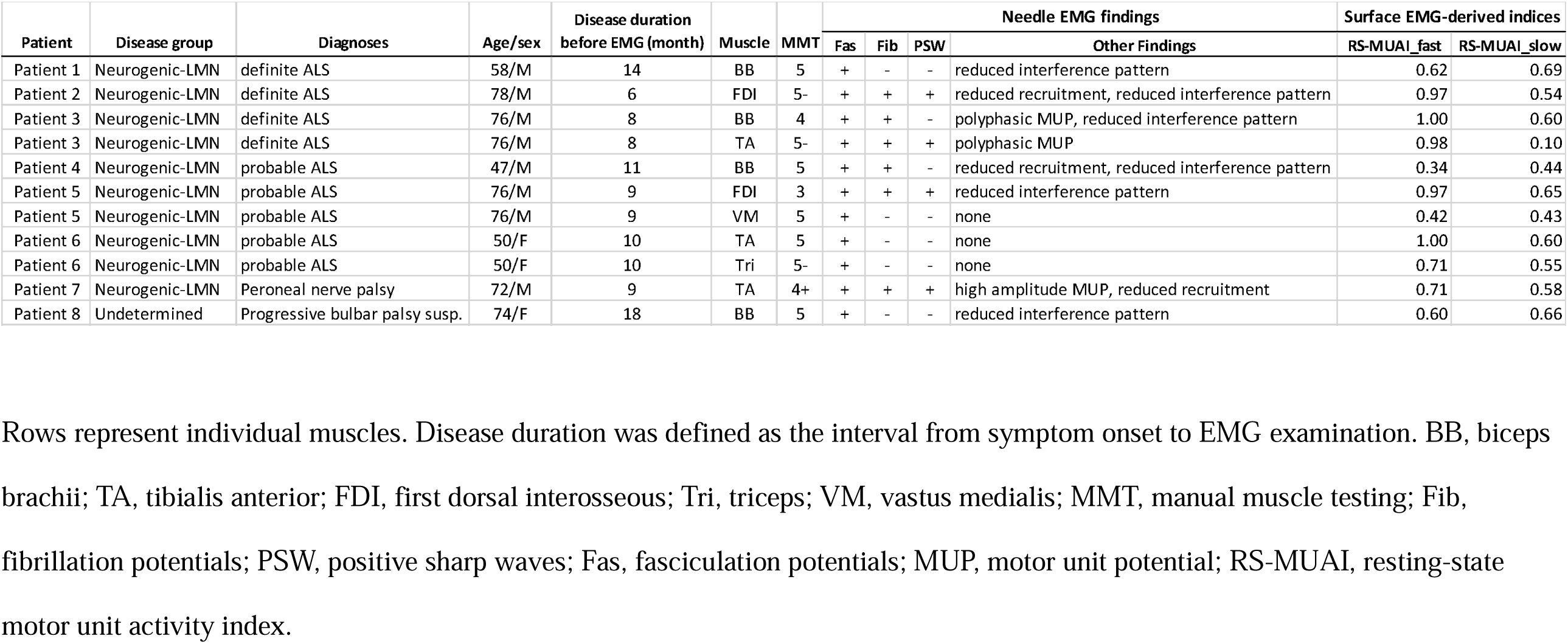
Needle EMG Findings and RS-MUAI Values in Fasciculation-positive Patients.

RS-MUAIs were compared between muscles with and without fasciculation potentials on nEMG (Fig. 3). RS-MUAI (slow) was significantly higher in the Fas-positive group (n = 11, median [IQR]: 0.58 [0.49–0.63]) than in the Fas-negative group (n = 56, 0.10 [0.00–0.29], p < 0.0001). Similarly, RS-MUAI (fast) was significantly higher in the Fas-positive group (median [IQR]: 0.71 [0.61–0.98]) than in the Fas-negative group (0.23 [0.12–0.40], p < 0.0001).

**Figure 3.**
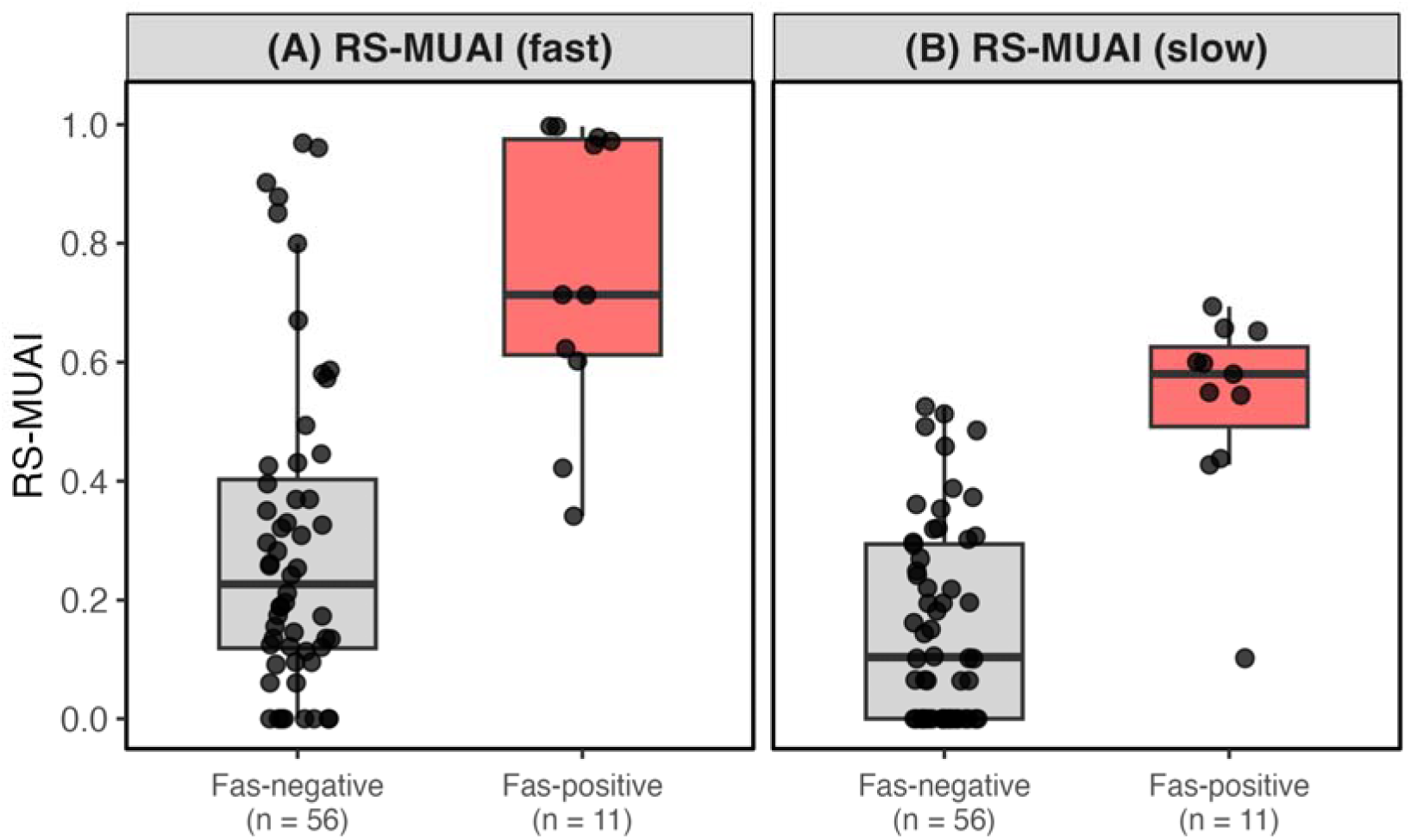
RS-MUAI (fast) and RS-MUAI (slow) according to fasciculation status. Box-and-whisker plots showing the distributions of (A) RS-MUAI (fast) and (B) RS-MUAI (slow) in muscles with fasciculation potentials on needle electromyography (Fas-positive, n = 11) and those without fasciculation potentials (Fas-negative, n = 56). Both RS-MUAI (fast) and RS-MUAI (slow) were significantly higher in Fas-positive muscles than in Fas-negative muscles (both p < 0.0001). Boxes indicate the interquartile range, horizontal lines indicate medians, whiskers indicate 1.5 times the interquartile range, and individual points represent muscle recordings.

In subgroup analyses based on diagnostic categories, significant differences were observed among groups for both RS-MUAI (slow) and RS-MUAI (fast) (Kruskal–Wallis test; Fig. 4). Post hoc analysis showed that the Neurogenic-LMN group had significantly higher values than the Myogenic and Normal groups for RS-MUAI (slow) (overall p < 0.0001; Neurogenic-LMN vs. Myogenic, p = 0.0015; Neurogenic-LMN vs. Normal, p < 0.0001). Similar patterns were observed for RS-MUAI (fast) (overall p = 0.0014; Neurogenic-LMN vs. Normal, p = 0.0009).

**Figure 4.**
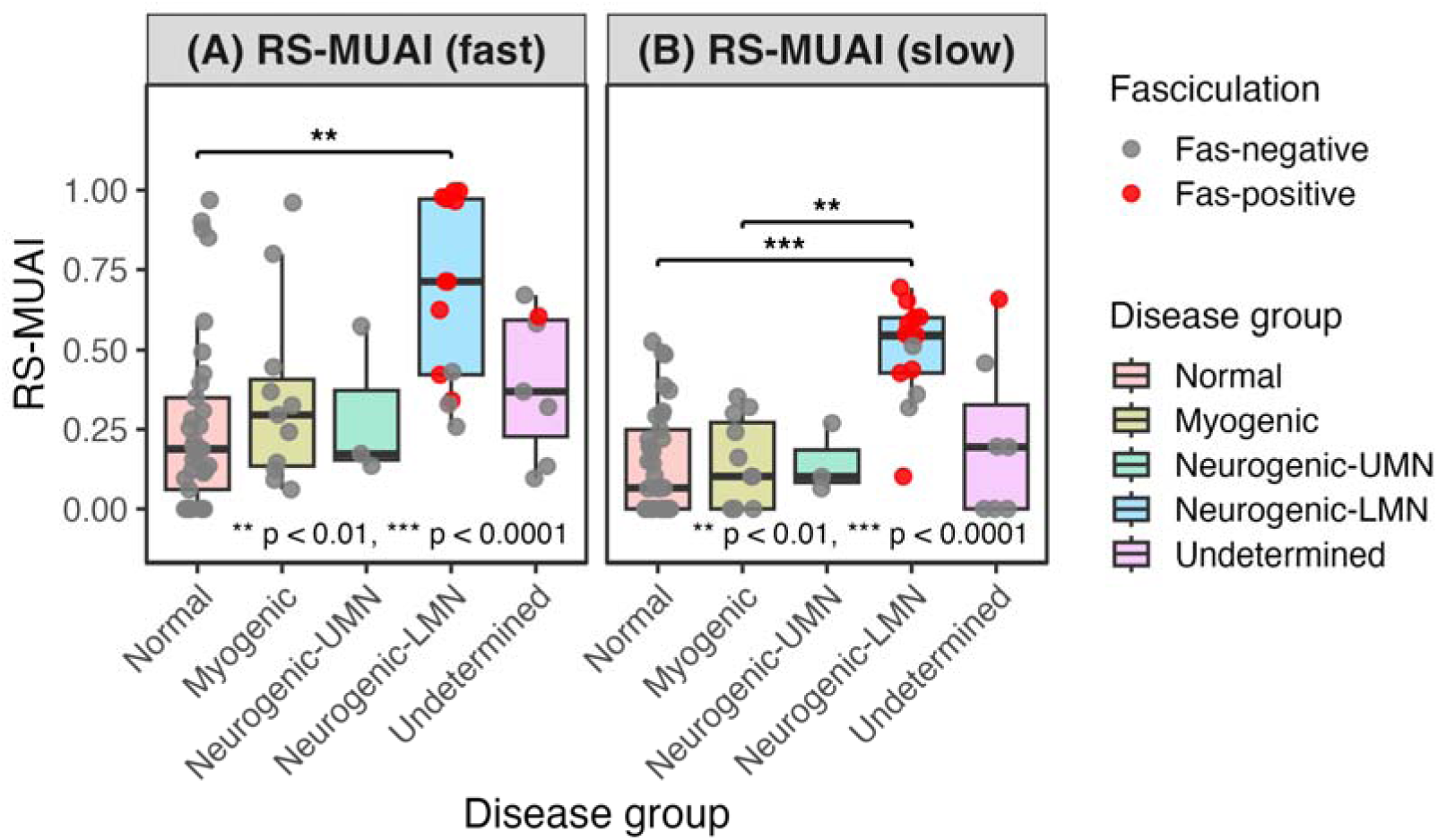
Comparison of RS-MUAIs by disease group. Box-and-whisker plots showing the distributions of (A) RS-MUAI (fast) and (B) RS-MUAI (slow) according to disease group: myogenic, neurogenic disorders with lower motor neuron involvement (Neurogenic-LMN), neurogenic disorders with upper motor neuron involvement without lower motor neuron involvement (Neurogenic-UMN), normal, and undetermined. Individual points represent muscle recordings and are colored according to fasciculation status on needle electromyography. Significant differences among disease groups were observed for both RS-MUAI (slow) and RS-MUAI (fast) using the Kruskal–Wallis test. The Neurogenic-LMN group showed the highest values for both indices, with significantly higher RS-MUAI (slow) than the myogenic and normal groups, and significantly higher RS-MUAI (fast) than the normal group in post hoc analyses. Boxes indicate the interquartile range, horizontal lines indicate medians, and whiskers indicate 1.5 times the interquartile range.

The GEE logistic regression model including RS-MUAI (fast) and RS-MUAI (slow) showed that both indices were significantly associated with the presence of fasciculation potentials on nEMG. RS-MUAI (slow) was associated with fasciculation potentials (odds ratio [OR] 5.10, 95% confidence interval [CI] 1.68–15.54, p = 0.0041), as was RS-MUAI (fast) (OR 2.38, 95% CI 1.17–4.78, p = 0.0162). When muscle region (upper vs. lower limb) was included in the GEE model, RS-MUAI (slow) remained significantly associated with fasciculation potentials, whereas RS-MUAI (fast) showed a positive but non-significant association. Muscle region itself was not significantly associated with fasciculation potentials. Similar results were observed in sensitivity analyses adjusting for age and sex, in which RS-MUAI (slow) remained significant, whereas RS-MUAI (fast) did not reach statistical significance.

Using each muscle as the unit of analysis and nEMG as the reference standard, we performed ROC analyses based on the predicted probabilities derived from the GEE model (Fig. 5). The MU-based sEMG indices showed good discriminative ability for identifying muscles with fasciculation potentials on nEMG.

**Figure 5.**
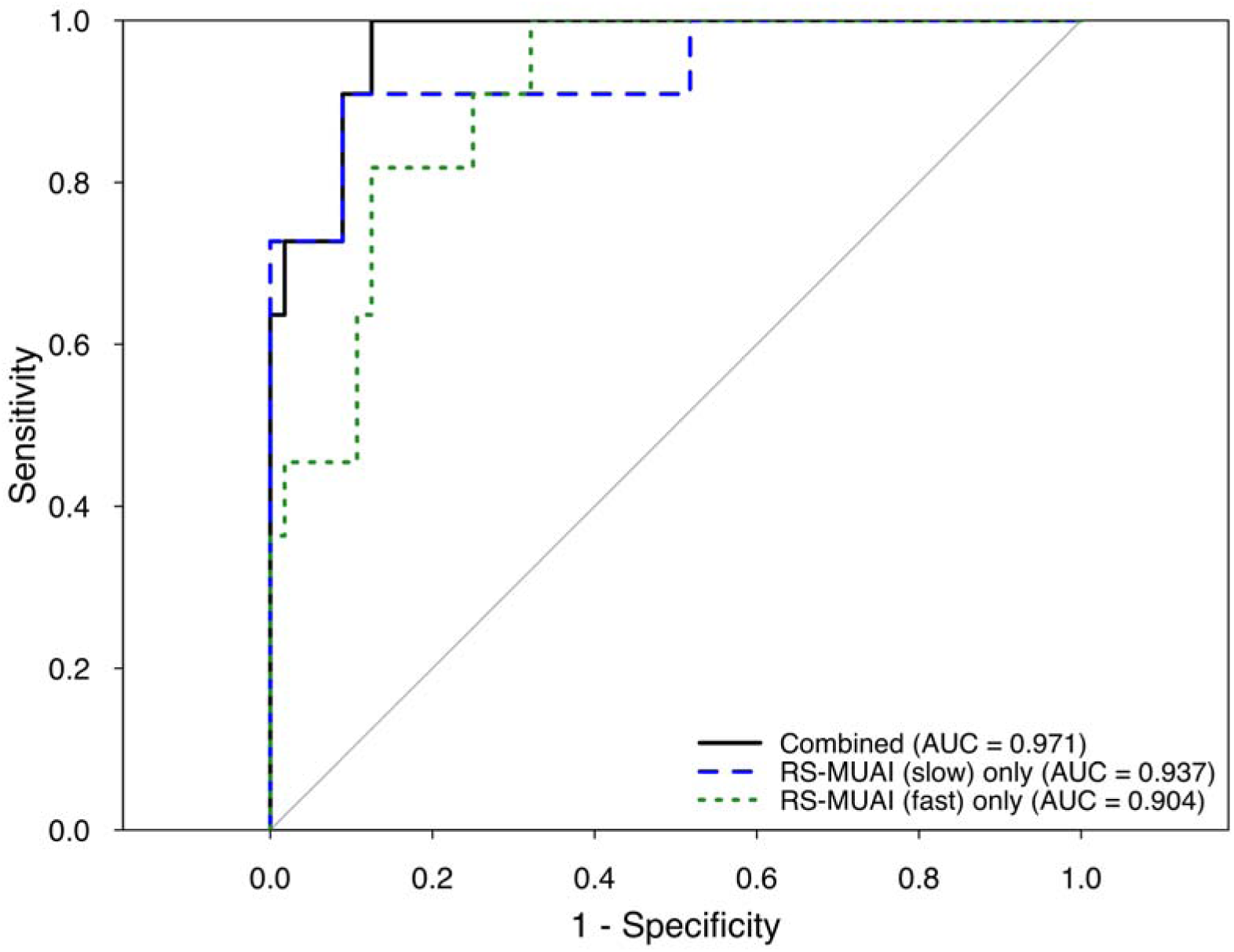
Receiver operating characteristic analysis for detecting fasciculation potentials on needle electromyography using RS-MUAI. Receiver operating characteristic (ROC) curves were generated to evaluate the ability of RS-MUAI (slow), RS-MUAI (fast), and their combined logistic model to identify muscles with fasciculation potentials on needle electromyography. The combined model showed the highest diagnostic performance, with an AUC of 0.971, compared with RS-MUAI (slow) alone (AUC, 0.937) and RS-MUAI (fast) alone (AUC, 0.904).

The area under the ROC curve (AUC) for the GEE model was 0.97 (95% CI, 0.94–1.00). At the optimal cut-off value of 0.12, sensitivity and specificity were 100% and 88%, respectively.

The AUCs for RS-MUAI (slow) and RS-MUAI (fast) alone were 0.94 (95% CI, 0.84–1.00) and 0.90 (95% CI, 0.82–0.98), respectively, both of which were lower than that of the combined GEE model, suggesting improved discriminative performance when the two indices were used in combination.

## 4. Discussion

In this study, the newly developed MU-based sEMG indices derived using the DEWCS method were significantly higher in fasciculation-positive muscles than in fasciculation-negative muscles. Using optimized cut-off values, these indices distinguished the two groups with high sensitivity and specificity in ROC analysis. These findings suggest that the DEWCS method-based analytical approach has potential as a screening tool for neuromuscular diseases, particularly in patients with amyotrophic lateral sclerosis (ALS).

Fasciculation potentials have particular clinical relevance in ALS because the Awaji criteria regard fasciculation potentials, when accompanied by chronic neurogenic changes, as electrophysiological evidence of lower motor neuron involvement (Costa et al., 2012, de Carvalho et al., 2008). Therefore, a non-invasive method capable of detecting fasciculation-related activity could provide clinically meaningful information before or alongside conventional nEMG.

Previous work on the application of sEMG to neuromuscular diagnosis has demonstrated the potential of non-invasive electrophysiological assessment. HD-sEMG studies have shown that non-invasive quantification of fasciculation potentials is feasible and diagnostically informative (Drost et al., 2007, Kleine et al., 2008, Tamborska et al., 2020). For example, Tamborska et al. reported that fasciculation frequency measured by HD-sEMG was markedly higher in ALS than in healthy and neurological controls, with high specificity for ALS prediction (Tamborska et al., 2020). However, HD-sEMG generally requires multi-electrode arrays, long recordings, and specialized analytical pipelines. In contrast, our approach uses a single bipolar recording channel, which may facilitate implementation in routine clinical EMG settings. In addition, the Clustering Index method, reported by Uesugi et al, has also demonstrated that single-channel sEMG can provide quantitative information useful for differentiating neurogenic and myopathic changes (Sonoo et al., 2021, Uesugi et al., 2011). Our method extends this concept by focusing on spontaneous MU-related activity at rest and by combining wavelet-based waveform extraction with component-specific MU activity indices.

A key feature of the present approach is that it does not rely solely on raw sEMG amplitude or visual identification of apparent fasciculations. Although obvious fasciculations may be visually recognizable on resting sEMG traces, visual inspection is subjective and may miss subtle or low-amplitude MU-related activity. In contrast, the DEWCS-based approach evaluates candidate events according to both MUAP-like waveform similarity and wavelet coefficient strength, and derives RS-MUAI from the ranked distribution of MR × WSS values, thereby providing a quantitative and reproducible measure of spontaneous MU-related activity. In the present study, RS-MUAI (slow) showed better diagnostic performance than RS-MUAI (fast) for identifying fasciculation-positive muscles, possibly because the duration and waveform characteristics of fasciculation-related activity were closer to the slow-type MU template. Nevertheless, the combined model using both RS-MUAI (slow) and RS-MUAI (fast) showed the highest diagnostic performance, suggesting that fast-type MU components may provide complementary information. Although the physiological interpretation of these component-specific indices requires further validation, this finding supports the value of decomposing resting sEMG activity into slow- and fast-type MU components.

Despite these advances, to our knowledge, no sEMG-based method has yet been widely adopted in routine clinical practice. The DEWCS-based approach may help bridge this gap because the recording procedure is similar to conventional sEMG, whereas the analytical output is expressed as objective quantitative indices. Clinically, this method may help avoid unnecessary nEMG examinations in patients with a low pre-test probability of abnormal spontaneous activity and may assist in selecting appropriate muscles for subsequent needle examination. In addition, because the method provides numerical outputs, it has potential as a biomarker for monitoring disease progression or treatment response, although longitudinal validation is required.

Importantly, the detection of fasciculation-related activity is not disease-specific. Fasciculations may occur in ALS as well as in benign fasciculation syndrome and other peripheral nerve disorders (Kleine et al., 2012, Mills, 2010). Therefore, RS-MUAI should not be interpreted as a stand-alone diagnostic marker for ALS, but rather as a non-invasive indicator of spontaneous MU-related activity that may guide further clinical and electrophysiological evaluation.

There are several limitations to our study. First, the sample size was small, and age and sex distribution differed among the diagnostic groups. Although the association between RS-MUAI (slow) and fasciculation potentials remained significant after adjustment for age and sex, the influence of demographic differences cannot be fully excluded. Second, because we needed to fine-tune the analytical method for detecting fasciculation potentials, a subset of the nEMG findings was not evaluated in a fully blinded manner during the early phase of the study. In addition, the cut-off values were both derived and evaluated within the same dataset, and their external validity has not been established; therefore, independent validation in a separate cohort is necessary. Third, there is a potential for selection bias, as the muscles selected for sEMG recording were chosen based on the nEMG findings. Finally, although we used generalized estimating equation models to account for within-subject correlation, residual non-independence at the subject level may still have influenced the results.

This prospective study demonstrated that sEMG combined with a novel analytical approach can identify muscles with fasciculation potentials on nEMG with promising accuracy. Further refinement of the analytical algorithm and validation in larger, multicenter cohorts are warranted to develop this method into a clinically applicable diagnostic tool.

## Author Contributions

**Takahiko Mukaino:** conceptualization, investigation, data curation, formal analysis, validation, funding acquisition, and writing – original draft.

**Hidetoshi Nagai:** conceptualization, methodology, software development, formal analysis, data curation, and writing – review & editing.

**Yuko Kobayakawa:** conceptualization, project administration, investigation, data curation, funding acquisition, and writing – review & editing.

**Senri Ko**: investigation, writing – review & editing.

**Kazunori Iwao**: investigation, writing – review & editing.

**Kotaro Iida**: investigation, writing – review & editing.

**Takashi Irie**: investigation, writing – review & editing.

**Saeko Inamizu**: investigation, writing – review & editing.

**Satoshi Nagata**: investigation, writing – review & editing.

**Eizo Tanaka**: investigation, writing – review & editing.

**Ryo Kurasawa**: investigation, writing – review & editing.

**Hajime Takeuchi**: investigation, writing – review & editing.

**Eri Miyazaki**: investigation, writing – review & editing.

**Noriko Isobe**: conceptualization, supervision, writing – review & editing.

**Hiroshi Shigeto**: conceptualization, supervision, writing – review & editing.

## Data Availability Statement

The data that support the findings of this study are available from the corresponding author upon reasonable request.

## Acknowledgements

We thank all the patients and volunteers involved in this study for their willingness and determination to participate. We also thank Drs. Koki Suezumi, Emiko Irie, and Mari Shinoda for their assistance with data acquisition. We are also grateful to Ms. Mika Umezaki for her assistance with scheduling.

## Declaration of generative AI and AI-assisted technologies in the manuscript preparation process

During the preparation of this work the authors used ChatGPT (OpenAI) and Claude (Anthropic) to assist with language editing, improving clarity, and refining the structure of the manuscript. After using this tool, the authors reviewed and edited the content as needed and take full responsibility for the content of the published article.

