## Supplementary Fig. 1 for "Non-invasive Detection of Fasciculation Using Surface EMG with a Wavelet-Based Analytical Method (DEWCS)"

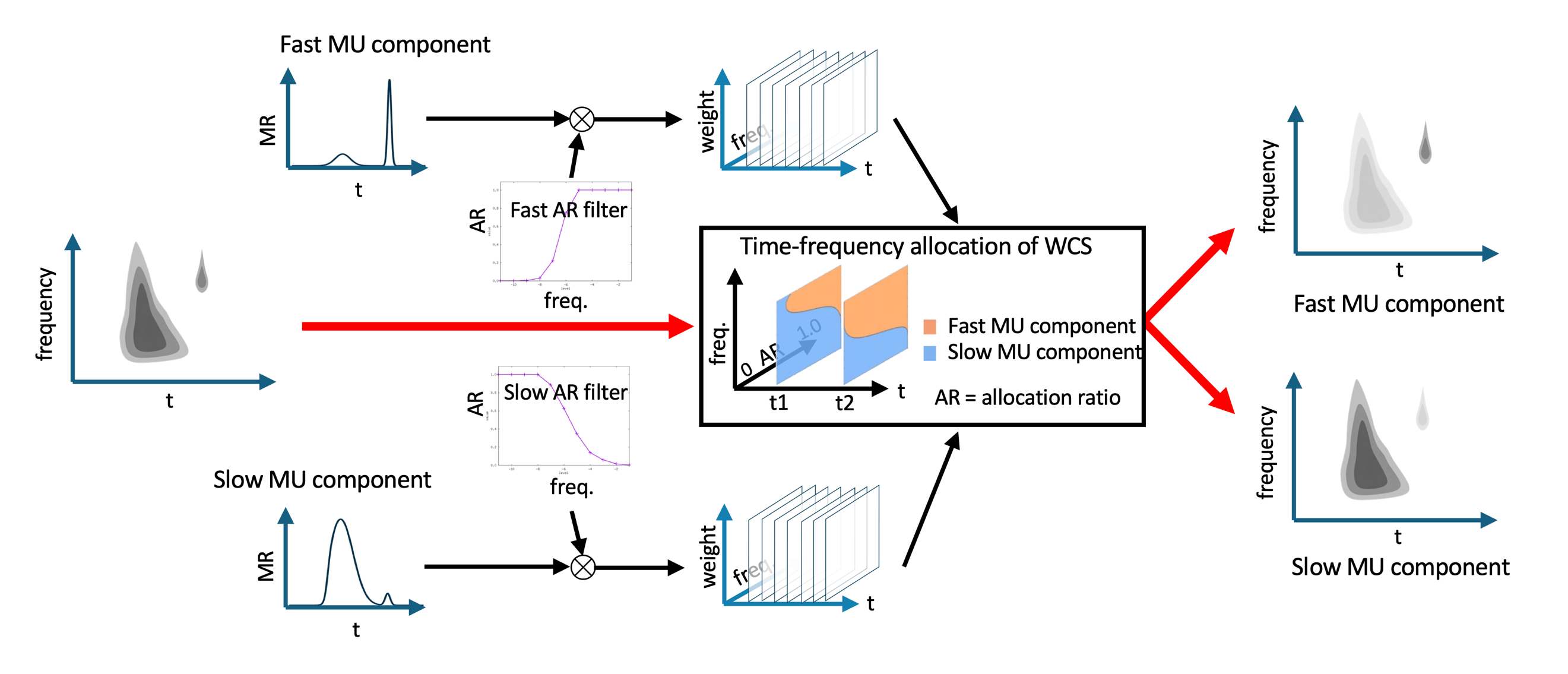

**Supplementary Figure 1. Schematic illustration of time–frequency allocation of a wavelet coefficient set into slow- and fast-type motor unit components.**
Matching ratios (MRs) were calculated separately for slow- and fast-type motor unit (MU) components. For time–frequency allocation of each wavelet coefficient set (WCS), the MR-based contribution of each MU component was further modulated by component-specific allocation ratio (AR) filters across frequency bands. The slow AR filter preferentially assigned lower-frequency coefficients to the slow-type MU component, whereas the fast AR filter preferentially assigned higher-frequency coefficients to the fast-type MU component. These frequency-dependent allocation weights were applied to the WCS at each time–frequency point, thereby generating slow- and fast-type MU component maps for subsequent calculation of component-specific WCS strength.
